# A Validated Measure of Rigidity in Parkinson’s Disease Using Alternating Finger Tapping on an Engineered Keyboard

**DOI:** 10.1101/2020.08.08.20170779

**Authors:** Megan H Trager, Kevin B Wilkins, Mandy Miller Koop, Helen Bronte-Stewart

## Abstract

**Introduction:** Reliable and accurate measures of rigidity have remained elusive in remote assessments of Parkinson’s disease (PD). This has severely limited the utility of telemedicine in the care and treatment of people with PD. It has also had a large negative impact on the scope of available outcomes, and on the costs, of multicenter clinical trials in PD. The goal of this study was to determine if quantitative measures from an engineered keyboard were sensitive and related to clinical measures of rigidity.

**Methods:** Sixteen participants with idiopathic PD, off antiparkinsonian medications, and eleven age-matched control participants performed a 30 second repetitive alternating finger tapping task on an engineered keyboard and were assessed with the Unified Parkinson’s Disease Rating Scale – motor (UPDRS-III).

**Results:** The speed of the key release was significantly slower in the PD compared to control cohorts (p <0.0001). In the PD cohort key release speed correlated with the lateralized upper extremity UPDRS III rigidity score (r = - 0.58, p < 0.0001), but not with the lateralized upper extremity tremor score (r = 0.14, p = 0.43).

**Conclusions:** This validated measure of rigidity complements our previous validation of temporal metrics of the repetitive alternating finger tapping task with the UPDRS III, bradykinesia and with the ability to quantify tremor, arrhythmicity and freezing episodes, and suggests that thirty seconds of alternating finger tapping on a portable engineered keyboard could transform the treatment of PD with telemedicine and the precision of multicenter clinical trials.

## Introduction

The diagnosis of Parkinson’s disease (PD) requires documentation of two motor signs among rigidity, bradykinesia, and/or resting tremor. Rigidity is assessed as increased resistance to passive range of movement of the limbs about the joints, requiring a face to face interaction between the PD individual and the neurologist. As telemedicine becomes a more common method of patient-physician interaction, the lack of a remotely accessed, reliable measure of rigidity limits the care of a person with PD.

The Unified Parkinson’s Disease Rating Scale (MDS-UPDRS) is the internationally accepted clinical scale to assess motor and non-motor aspects of PD [1]. For the MDS-UPDRS III (motor subscale) the examiner rates motor function on an integer scale from zero (normal) to four (incapacity to perform the task). There are several challenges of the MDS-UPDRS III: it is subjective, there is significant inter- and intra-rater unreliability, and it requires face-to-face evaluation of the person with PD. The MDS-UPDRS III is required for any clinical trial in PD. It is video-recorded and then assessed by a “blinded rater” to standardize outcomes. Unlike bradykinesia or tremor, rigidity cannot be assessed from a video and must be dropped from the ratings. These shortcomings have had a negative impact on the scope of the available outcomes and have increased the costs of performing pivotal multicenter clinical trials.

This report will demonstrate the solution to this unmet need with the first validated, quantitative measure of rigidity in PD, using a repetitive alternating finger tapping task (RAFT) on an engineered keyboard that measures key displacement with submillimeter accuracy. The RAFT task was originally chosen as it is a well learned motor task but would elicit more fine motor control abnormalities in PD than single digit tapping [2-6]. Performance of RAFT requires the passive release of one finger as the other finger actively presses the key. We hypothesized that the speed of the release of the key would be correlated with the examiner’s assessment of rigidity from the UPDRS III.

## Methods

### Human Subjects

Sixteen subjects (twelve men) with idiopathic PD and 11 controls (five men) were recruited from the Stanford Movement Disorders Center and consented to participate in the study, which was approved by the Stanford Institutional Review Board. A fellowship-trained Movement Disorders specialist confirmed the diagnosis in each subject. Each side of the body was treated as a separate data point.

### Experimental Protocol

PD individuals were assessed using QDG and the UPDRS III in the off-medication state: long acting and short acting dopaminergic medication were held for >24 and >12 hours respectively prior to testing. Individuals sat on an armless chair with the elbow flexed at 90 degrees and the wrist resting on a pad at the same level as the keys of a customized engineered keyboard [2]. They placed the index and middle finger on adjacent keys. The instructions were to tap each key in an alternating pattern, as fast and as regularly as possible for 30 seconds, starting and stopping only when they heard an auditory cue. They were instructed to attempt to press and release the keys completely. No external pacing or cueing was provided. Subjects performed the tasks without visual (eyes closed) or auditory feedback [2, 6-8]. Each subject had a short period of practice before the test began, and all trials were videotaped; the video of the fingers was time-stamped and synchronized with the data. The corresponding upper extremity rigidity score (maximum score of 4) and postural and resting tremor scores (maximum score of 8) of the UPDRS III were used.

### Data acquisition and analysis

The keyboard produced a voltage signal that was proportional to the displacement of the key. The key displacement was linearly related to the output voltage signal with a resolution of 62.5 um per 40 mV. For key displacements less than 9 mm, the keyboard operated in a linear zone. Near the base of the key displacement, the key reached a compliant mechanical stop and when the finger continued to press done, the additional displacement was non-linearly related to the output voltage signal, Figure 1A [3].

**Figure 1.**
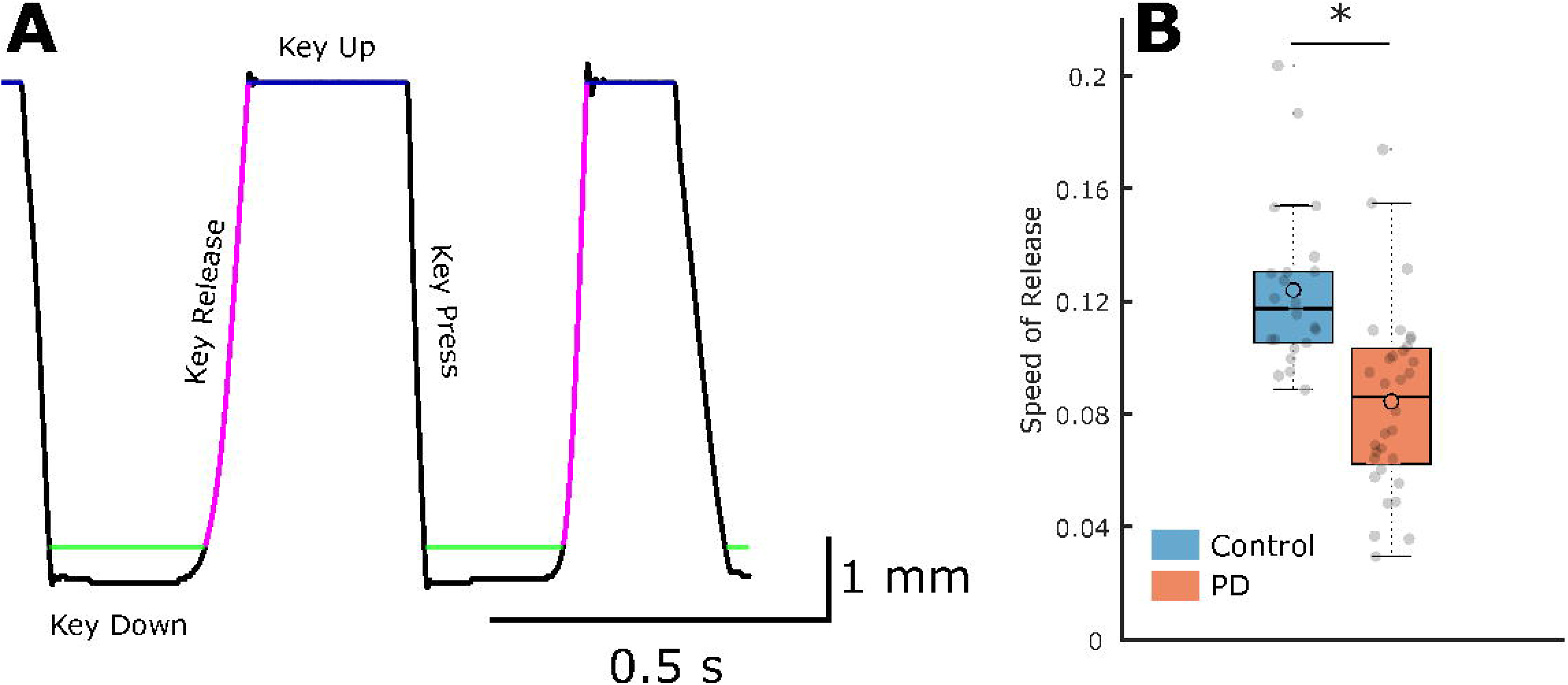
**A**. Example of several cycles of the finger tapping task from one finger in an individual, demonstrating the downward and upward motion of the key (key press and key release), respectively. Key Down and Key Up represent the furthest extent of the key press and the phase when the finger has released the key, respectively. The green line marks the end of the linear zone^7^. The speed of key release is calculated from the slope of the key release (magenta line). **B**. Boxplot showing the speed of release in PD individuals and age matched controls. The speed of release in PD patients (orange) was significantly slower than those of control subjects (blue). * P < 0.0001.

A customized detection algorithm, written in MATLAB, was used to determine specific states in the cycle of finger tapping movement, Figure 1A.

The amplitude of each key strike was calculated for key displacements in the linear zone. The speed of the key release was defined as the ratio of the amplitude of the key release for each strike to the time between key down and key up and was the average of the total key releases for both fingers.

### Statistics

A linear mixed model was used to compare the speed of key release between the control and PD cohorts, looking at group differences in speed of release while controlling for sex. A Spearman correlation was used to assess the relationship between the average speed of the release and the UPDRS scores for rigidity and tremor.

## Results

The mean ± SD age of the PD group was 68.9 + 8.7 years with a disease duration of 8.2 + 5.5 years. The mean off therapy UPDRS III score ± SD was 35.5 ± 11.3, and the lateralized scores for the upper extremities were 10.2 ± 4.7 and 6.2 ± 3.2 in the more affected and less affected sides, respectively. The mean ± SD age of control group was 63.2 + 6.6 years.

The speed of key release speed was significantly faster in the control compared to the PD individuals (t(51) = 5.54, p = 1.05e-6), Fig.1B. Within the PD cohort the speed of key release was significantly correlated with the same upper extremity UPDRS III rigidity score (r = - 0.58, p < 0.0001), Figure 2A. However, there was no significant correlation with the speed of key release and the UPDRS III lateralized upper extremity tremor score (r = 0.14, p = 0.43), Fig. 2B.

**Figure 2.**
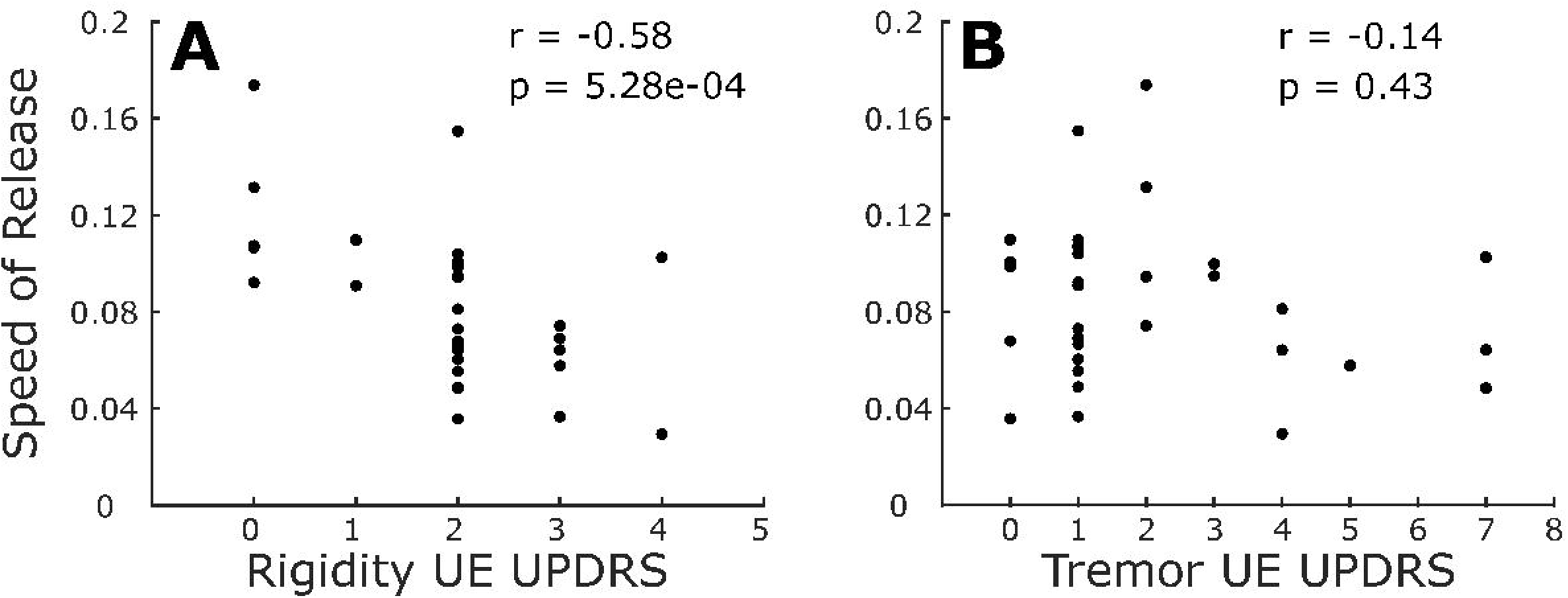
Correlation between release speed and lateralized UPDRS III items, **A:** rigidity, and **B:** tremor.

## Discussion

This study has demonstrated that the speed of the release of the key during a thirty second repetitive alternating finger tapping task (RAFT) on an engineered keyboard was a robust quantitative measure of upper extremity rigidity in individuals with PD, that was validated with the UPDRS III subscore of rigidity; the lower the speed of release the greater the rigidity. The speed of key release in the PD cohort was significantly slower than that in age-matched healthy controls. To our knowledge, this is the first demonstration of a validated, quantitative measure of rigidity using a simple thirty second alternating finger tapping task on an engineered keyboard, which accurately measures amplitude of the key displacement.

### The release phase of repetitive alternating finger tapping is a correlate of passive range of motion

Rigidity is experienced by the examiner as a “plastic” or “lead pipe” resistance to her/his passive movement of the limb around a joint. The RAFT task is a well learned movement, whose active component is to press one key down, while the other is released. We hypothesized that the key release is a form of passive movement as the finger is pushed up by the spring in the key. If there is rigidity in the upper extremity the passive release may be slower.

RAFT on the engineered keyboard provides an objective, accurate, and quantifiable measure of rigidity; it requires technology that can measure the amplitude of displacement of the keys accurately. We originally introduced the technology of Quantitative DigitoGraphy (QDG) using the RAFT task on a musical instrument digital interface (MIDI) keyboard [6]. The original report demonstrated that temporal measures such as the frequency of tapping and the duration of finger strike could differentiate people with PD from healthy controls, and people with PD when off compared to on dopaminergic medication [6]. QDG in the PD cohort identified tremor and freezing episodes during RAFT. We subsequently validated the temporal metrics of the RAFT task on a MIDI keyboard with the overall UPDRS III score and bradykinesia [7] and demonstrated that disorders of fine motor control, reflected in the RAFT, were sensitive to motor control abnormalities in very early stage PD [8, 9].

The major limitation of the MIDI keyboard was that it did not accurately measure the amplitude of the key displacement. To address this limitation, a prototype of an engineered keyboard was developed, which measured the amplitude of key displacement using optical sensors, and with which we demonstrated the superiority of the RAFT task to a single digit tapping task for the measurement of arrhythmicity of tapping in PD [2]. In this study, we demonstrate for the first time that the speed of key release is a validated, robust quantitative measure of rigidity in PD.

Previously, technologies such as electromyography (EMG), inertial measuring units (IMUs), potentiometers, and electrically driven servomotors have provided valid objective metrics of rigidity [10-12]. These technologies offer limited utility in telemedicine care because the size and/or complexity of the device pose significant challenges for patients with PD to perform self-administered tests, and they have not been shown to measure other cardinal motor signs, as we have demonstrated with QDG.

### The absence of a remote, reliable measure of rigidity is a major limitation for telemedicine and multicenter clinical trials in Parkinson’s disease

The challenges of the UPDRS III have significantly impacted the neurologist’s ability to accurately assess and treat the person with PD using telemedicine. Across the world, telemedicine visits have exploded in demand during the coronavirus disease of 2019 (COVID-19) pandemic. PD is a chronic, progressive disease, and requires frequent monitoring to optimize and titrate therapies for effective treatment. Hence, the utility of a remotely assessed validated measure of rigidity, using a simple thirty second RAFT task will improve patient care for PD patients and other patient populations as well, via telemedicine.

In multicenter clinical trials the UPDRS III is recorded and then assessed via video by a “blinded rater” to standardize outcomes, due to the poor inter-rater reliability of scoring. As rigidity cannot be assessed from a video it must be dropped from the assessments. This has a negative impact on the scope of the available outcomes for any PD therapeutic. The need for a face-to-face, examiner-patient interaction, the subjective nature, and lack of inter-rater reliability of the UPDRS III have resulted in large increases in the costs of performing multicenter clinical trials, which require increased sample sizes needed for statistical significance.

The challenges to telemedicine care and clinical trials can now be answered using RAFT on an engineered keyboard, which provides an objective, quantifiable measure of rigidity and other motor signs in PD. The engineered keyboard is a portable tool, which provides comprehensive measures of motor control in PD (rigidity, bradykinesia, tremor, freezing episodes) and has been validated with the overall UPDRS III, the use of which will transform remote care of the person with PD and the scope and cost of clinical trials [6-9]. It is a sensitive tool to detect motor disability in very early stages of disease [8, 9] and potentially could be used to predict the onset of PD in susceptible individuals.

## Conclusion

Rigidity assessment in PD requires an in-person examination and cannot be rated from a video recording of the UPDRS III. This is a critical unmet need and challenge for both telemedicine and multicenter clinical trials. In this study we validated an objective measure of rigidity in PD using a thirty second RAFT task on an engineered keyboard, which measures key displacement with submillimeter accuracy. The speed of the release of the key was significantly slower in the PD compared to control cohorts; it was strongly correlated with rigidity and was not related to tremor. The addition of a validated quantitative metric for rigidity allows RAFT on an engineered keyboard to serve as a remote reliable quantitative measure of all cardinal motor signs of PD, that will enhance the precision of telemedicine and will improve the accuracy and reduce the cost of multicenter clinical trials. QDG metrics can detect motor disability in very early stages of disease and potentially could be used to predict the onset of PD in susceptible individuals.

## Data Availability

Data is provided in the manuscript text. Data sets can be requested from the authors.

